# Use of Metagenomic Microbial Plasma Cell-Free DNA Next-Generation Sequencing Assay in Outpatient Rheumatology Practice

**DOI:** 10.1101/2024.09.09.24313356

**Authors:** Rachel Jenkins, Matthew Samec, Courtney Arment, Kenneth J. Warrington, John M. Davis, Matthew J. Koster

**Author notes:** CORRESPONDING AUTHOR: Rachel Jenkins, M.D., Department of Internal Medicine Mayo Clinic, 200 First Street SW Rochester, MN, USA 55905. **Financial Disclosure**: The was no funding for this study and no author has any financial disclosures affiliated with this work.

## Abstract

**Objectives:** To assess the utility of a metagenomic microbial plasma cell-free DNA next-generation sequencing assay (Karius Test^TM^; KT) in the evaluation of patients in an outpatient rheumatology practice.

**Methods:** All patients with a KT ordered and obtained by a rheumatology provider in the outpatient setting from 1 January 2020 through 31 December 2022 were retrospectively identified. Demographic, clinical, laboratory, radiologic, histopathology, and microbial studies were abstracted. Indication for KT testing was categorized. KT results were defined based on positive result and clinical relevance regarding the symptoms under investigation at the time of the rheumatologic investigation. Review of cases three months after KT was undertaken to determine clinical outcome.

**Results:** 150 patients with a KT were included (53% female, mean age 52 years). The reason for KT was evaluation of atypical presentation of rheumatic disease (80%), assessing flare versus infection in patient on immunosuppression (16.7%), and fever of unknown origin (3.3%). 24 (16%) KTs were positive, 6 of which were considered clinically relevant and altered the final diagnosis and treatment. Of the 126 negative KTs, 5 (4%) were found to have a clinically relevant infection by conventional testing methodologies.

**Conclusions:** In this large retrospective cohort study, the most frequent reason for KT utilization was an atypical presentation of rheumatic disease. 25% of positive KTs altered the final diagnosis and treatment, whereas a false-negative rate of 4% was observed. KT has utility in the outpatient rheumatology assessment. Further delineation of which patients are best suited for KT testing remains to be defined.

**Key messages:**

1. Use of microbial cell free DNA testing assisted in differentiating between atypical infection and rheumatologic presentation
2. Further studies are needed to define the most optimal patient selection for maximizing mcfDNA technologies in the rheumatology outpatient setting

## Introduction

For all patients presenting to the rheumatology clinic, clinicians are tasked with determining if a patient’s presentation is due to an autoimmune condition. This can be difficult as inflammatory syndromes often have overlapping clinical features with a wide spectrum of diseases, including indolent infection. For example, it is well known that infective endocarditis is associated with ANCA serology positivity and features of vasculitis, [1] and the diagnosis of rheumatoid arthritis confers a higher infection risk both due to its pathophysiology and treatments [2]. These overlapping clinical features are particularly important for patients where infection must be excluded prior to considering initiation or escalation of immunosuppression. Metagenomic next-generation sequencing (mNGS) of microbial cell-free DNA (mcfDNA) allows for non-invasive detection of fragments of pathogen (bacteria, DNA virus, fungus, and parasite) DNA deposited into the blood from distant infection sites and does not rely on organism propagation on a growth medium[3, 4].

The Karius test (KT) is a commercially available mNGS mcfDNA technology that can evaluate over 1000 pathogens. Its utility has already been described for non-invasively identifying musculoskeletal infections, infective endocarditis, community-acquired and post-viral pneumonia [4–6]. Efficacy of its use has also been evaluated in immunocompromised populations with a focus on patients with hematologic malignancy, bone marrow transplant, and solid organ transplant as well as those recently treated with antibiotic therapy [3, 4, 7, 8]. Assessment of additive diagnostic value of mNGS mcfDNA testing has been investigated in immunocompetent patients presenting with fever of unknown origin [9], but there is a lack of data regarding the benefit of KT among patients with atypical presentations of rheumatic disease and limited information in patients with autoimmune diseases on conventional or biologic immunosuppressive medications. Furthermore, the majority of studies to date have focused on mNGS testing during inpatient hospitalization, therefore the relevance of outpatient testing remains unexplored. The aim of this study was to describe the clinical utility of KT in a tertiary, outpatient rheumatology practice.

## Patients & Methods

We conducted a retrospective chart review of all patients for whom a KT was obtained in the outpatient clinical setting by a provider in the Division of Rheumatology at Mayo Clinic, Rochester between 1 January 2020 and 31 December 2022. This study was deemed Exempt by the institutional IRB. Demographic, clinical, laboratory, radiographic, and histologic data abstracted from the electronic medical record (EMR) included gender, age, reason for testing, presenting symptoms, white blood cell count, hemoglobin, creatinine, immunologic serologies, inflammatory markers, current medications (disease-modifying anti-rheumatic drugs [DMARDs], biologics, glucocorticoids, and antibiotics within past 30 days), results of the KT and other infectious workup, and clinical outcome (changes to immunosuppression versus infection diagnosis). Reason for testing was categorized as one of the following: (1) to assess fever of unknown origin (FUO), (2) to assess an atypical rheumatic disease (RD) presentation, or (3) to assess for RD flare versus infection in the setting of immunosuppression. The KT result was determined to be one of the following: (1) positive (≥1 pathogen identified) and clinically relevant, meaning the pathogen was deemed to be associated with presentation for which the KT was obtained, and anti-microbial therapy targeting the positive KT result was initiated, (2) positive and clinically irrelevant, meaning the pathogen was deemed to not be associated with the clinical presentation or symptoms for which rheumatologic evaluation was requested [e.g. KT positive for Escherichia coli in setting of urinary tract infection unrelated to symptoms prompting rheumatology consultation], or (3) negative. The medical charts were reviewed up to three months after the KT to evaluate the clinical outcome, including initiation or enhancement of immunosuppression and identification of infectious organism via alternative testing methods.

## Results

One hundred and fifty patients were identified as having a KT obtained in the outpatient rheumatology clinic during the study period. Baseline characteristics, including sex, age, reason for testing, and other laboratory values obtained at the time of testing, are included in Table 1. Of all patients for which a KT was obtained, 53% were female, and the mean age was 52 years. The majority of KTs (80%) were obtained for assessment of an atypical RD presentation with the remainder being tested for assessing RD flare versus infection (16.7%) and fever of unknown origin (3.3%).

**Table 1.**
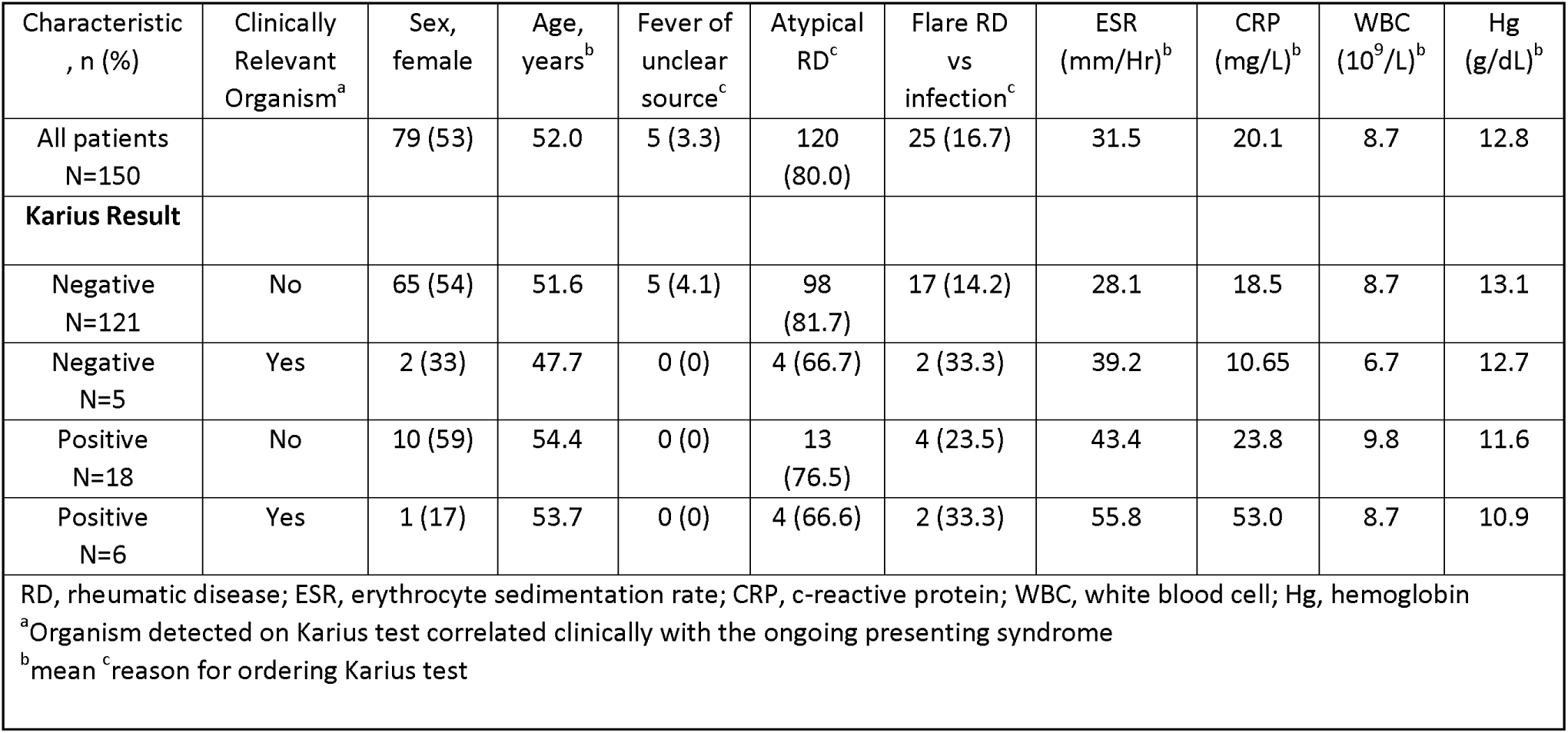
Baseline characteristics of Karius testing groups.

Of the 150 KTs completed, 24 (16%) were positive. Twenty-five percent of the positive tests (6/24) were considered clinically relevant and altered final diagnosis and treatment. The remainder were deemed clinically irrelevant in regard to the symptoms present for which the rheumatologic evaluation was taking place. Infectious disease consultations were obtained in all circumstances in which clinical relevance was unclear. Table 2 describes the determined clinical relevance of all KT-identified organisms. Eight percent (2/25) of KTs obtained to evaluate for RD flare vs infection and 4/120 (3%) of those obtained to evaluate atypical RD were deemed positive, clinically relevant results. No tests obtained for FUO were deemed positive, clinically relevant results. Patients with positive, clinically relevant results tended to have higher erythrocyte sedimentation rate (ESR) and C-reactive protein (CRP) at 55.8 mm/hr and 53.0 mg/L, respectively. Whereas the ESR and CRP for patients with negative KT, but for whom an infectious organism was identified by another methodology were 44.3 mm/hr and 9.2 mg/L, respectively. Of the patients with a positive KT, 20/24 (83%), including 4/6 (67%) patients with a positive and clinically relevant organism identified, were on immunosuppression (DMARDs, biologic, or glucocorticoids) at the time of testing.

**Table 2.**
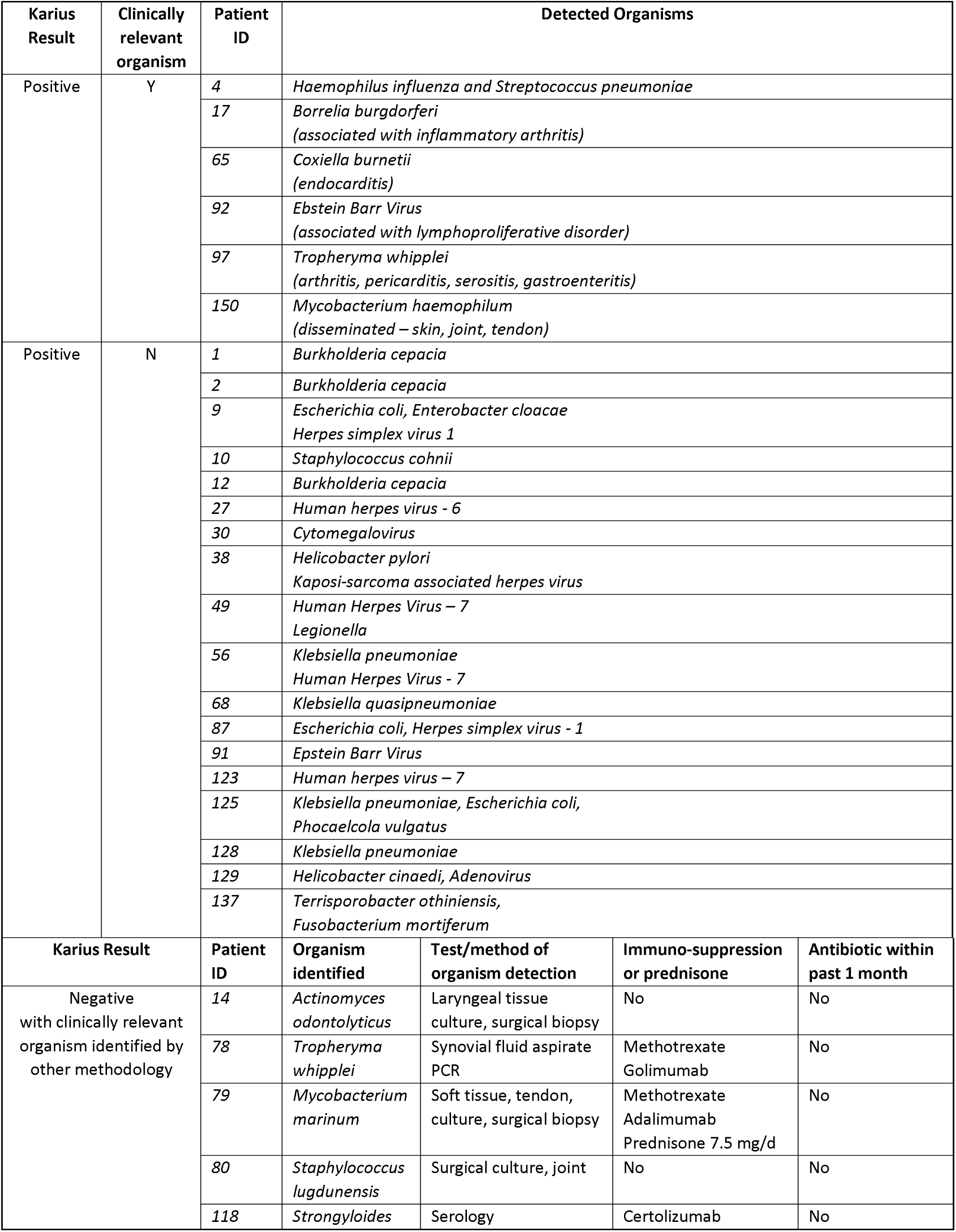
Karius test result and organisms identified.

| Karius Result | Clinically relevant organism | Patient ID | Detected Organisms |  |  |  |
| --- | --- | --- | --- | --- | --- | --- |
| Positive | Y | 4 | *Haemophilus influenza* and *Streptococcus pneumoniae* |  |  |  |
|  |  | 17 | *Borrelia burgdorferi*<br>(associated with inflammatory arthritis) |  |  |  |
|  |  | 65 | *Coxiella burnetii*<br>(endocarditis) |  |  |  |
|  |  | 92 | *Epstein Barr Virus*<br>(associated with lymphoproliferative disorder) |  |  |  |
|  |  | 97 | *Tropheryma whipplei*<br>(arthritis, pericarditis, serositis, gastroenteritis) |  |  |  |
|  |  | 150 | *Mycobacterium haemophilum*<br>(disseminated – skin, joint, tendon) |  |  |  |
| Positive | N | 1 | *Burkholderia cepacia* |  |  |  |
|  |  | 2 | *Burkholderia cepacia* |  |  |  |
|  |  | 9 | *Escherichia coli*, *Enterobacter cloacae*<br>*Herpes simplex virus 1* |  |  |  |
|  |  | 10 | *Staphylococcus cohnii* |  |  |  |
|  |  | 12 | *Burkholderia cepacia* |  |  |  |
|  |  | 27 | *Human herpes virus - 6* |  |  |  |
|  |  | 30 | *Cytomegalovirus* |  |  |  |
|  |  | 38 | *Helicobacter pylori*<br>*Kaposi-sarcoma associated herpes virus* |  |  |  |
|  |  | 49 | *Human Herpes Virus – 7*<br>*Legionella* |  |  |  |
|  |  | 56 | *Klebsiella pneumoniae*<br>*Human Herpes Virus - 7* |  |  |  |
|  |  | 68 | *Klebsiella quasipneumoniae* |  |  |  |
|  |  | 87 | *Escherichia coli*, *Herpes simplex virus - 1* |  |  |  |
|  |  | 91 | *Epstein Barr Virus* |  |  |  |
|  |  | 123 | *Human herpes virus – 7* |  |  |  |
|  |  | 125 | *Klebsiella pneumoniae*, *Escherichia coli*,<br>*Phocaelcola vulgatus* |  |  |  |
|  |  | 128 | *Klebsiella pneumoniae* |  |  |  |
|  |  | 129 | *Helicobacter cinaedi*, *Adenovirus* |  |  |  |
|  |  | 137 | *Terrisporobacter othiniensis*,<br>*Fusobacterium mortiferum* |  |  |  |
| Karius Result |  | Patient ID | Organism identified | Test/method of organism detection | Immuno-suppression or prednisone | Antibiotic within past 1 month |
| Negative<br>with clinically relevant<br>organism identified by<br>other methodology |  | 14 | *Actinomyces odontolyticus* | Laryngeal tissue culture, surgical biopsy | No | No |
|  |  | 78 | *Tropheryma whipplei* | Synovial fluid aspirate PCR | Methotrexate<br>Golimumab | No |
|  |  | 79 | *Mycobacterium marinum* | Soft tissue, tendon, culture, surgical biopsy | Methotrexate<br>Adalimumab<br>Prednisone 7.5 mg/d | No |
|  |  | 80 | *Staphylococcus lugdunensis* | Surgical culture, joint | No | No |
|  |  | 118 | *Strongyloides* | Serology | Certolizumab | No |

Clinically relevant organisms (Table 2) identified by a KT, included organisms such as Borrelia burgdorferi, Tropheryma whipplei, and Mycobacterium haemophilum. The symptoms associated with these organisms were consistent with the clinical features that resulted in request for rheumatologic consultation. The amount of time taken to identify Mycobacterium haemophilum (patient 150) by KT was 48 hours, whereas confirmation of this organism by conventional microbial testing methods took six weeks by blood culture, synovial fluid aspirate, and tissue culture [Figure 1] Clinically irrelevant organisms (Table 2) identified by a KT included those that may confer lifelong infection once exposed without clinical relevance, such as Human Herpes Virus – 6, Human Herpes Virus-7, Cytomegalovirus, and Epstein-Barr Virus. While organisms seen on KT may have been pathogenic, they were not attributed to the symptomatology for which the KT was obtained. Most frequent among the positive but clinically irrelevant organisms were urinary and pulmonary microbes such as Escherichia coli, Klebsiella pneumoniae and Burkholderia cepacia.

**Figure 1.**
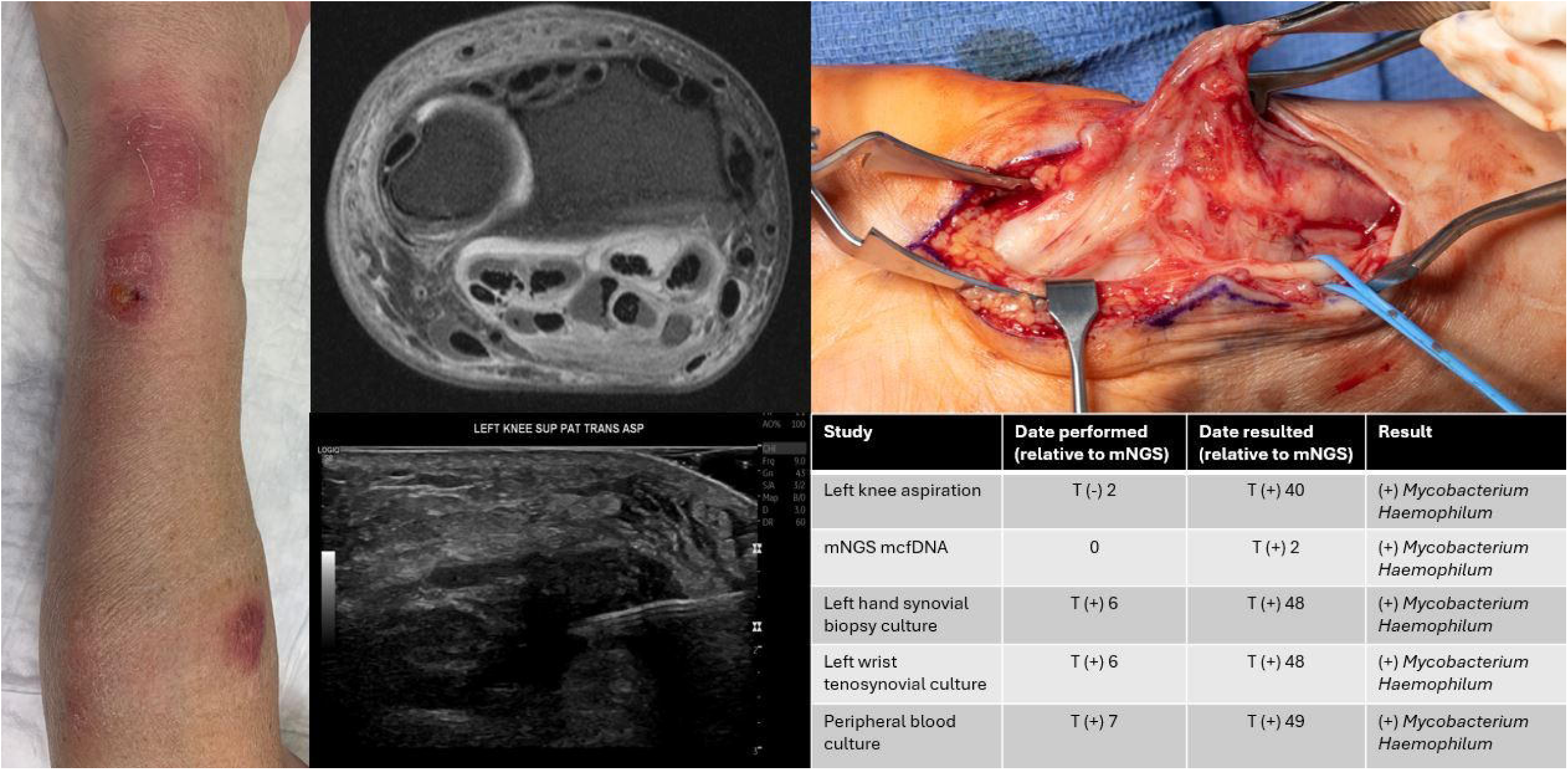
Erythematous, indurated, nodular lesions over the wrist and distal and proximal right forearm (left). Contrast enhanced MRI with diffuse T2 hyperintensity and tenosynovial thickening in the right wrist flexor tendons (axial view; Middle Upper). Ultrasound guided aspiration of left knee effusion with notable gray-scale synovitis (transverse view; middle lower). Right forearm, volar intraoperative view, demonstrating tenosynovial thickening and rice bodies (right upper). Timing of studies and results relative to mNGS mcfDNA collection date (right, lower).

Out of the 126 (84%) negative KTs, 5 (4%) were subsequently found to have a clinically relevant infection identified by other methodology. Eighty percent (4/5) of these patients required invasive testing, such as surgical biopsy/culture or synovial fluid aspirate PCR to identify the pathogenic organism. Three (60%) of these patients were on immunosuppressive agents at the time of diagnosis, of which all three were receiving a tumor necrosis factor (TNF) alpha inhibitor alone or in combination. The organism identified, methodology by which the organism was identified, and immunosuppressive regimen at the time of negative KT are described in Table 2.

Among the patients with a negative KT and no infectious organism identified by other methodology, 60/121 (49.5%) were on immunosuppression at time of obtaining the KT. Among patients with a negative KT 44% (55/126) were started on immunosuppression and 9% (11/126) escalated immunosuppression after the negative KT. None of the patients initiating or escalating immunosuppression after negative KT were identified to have an infection associated with the initial presentation in the subsequent 3 months following the KT test.

## Discussion

Given infections can mimic rheumatic disease and patients with known, established rheumatic diseases on immunosuppression are at increased risk for opportunistic infections, identification of infection as the cause of symptoms is of critical importance in the evaluation of patients in a rheumatology practice. Metagenomic sequencing of mcfDNA provides a method for non-invasive testing with broad coverage of bacterial, mycobacterial, fungal, and viral organisms with fast turnaround. This study is the first to describe use of mNGS of mcfDNA by KT in a large, tertiary referral outpatient rheumatology setting.

Metagenomic mcfDNA testing performed in outpatients presenting for rheumatologic evaluation resulted in positive detection of a pathogen in 24 (16%), of which 25% of cases were felt to be clinically relevant. Our series has a notably lower pathogen detection rate than other cohorts using similar mcfDNA technology where rates of pathogens in ≥ 1 taxon are 38-71% [6, 8, 10, 11]. The discrepancy between our cohort and others is likely due to inherent differences in the risk profiles and testing indications. Whereas unexplained fever, sepsis, or leukocytosis were the predominant reasons for mcfDNA testing during hospitalization evaluation [6, 10], atypical presentation of rheumatic disease accounted for 80% of testing while FUO was the least common of the three main indications for testing in our outpatient cohort (3%). In addition, the majority of patients evaluated with mcfDNA in reported studies represent immunocompromised patients with hematologic malignancies, hematopoietic stem cell transplant, solid organ transplant, or primary immunodeficiency [6–8, 10–13]. Limited information is available on the testing characteristics among patients with autoimmune disease. Among studies that have evaluated mcfDNA testing in individuals with autoimmune disease, such patients have represented only 4-7% of the total cohort [10, 14].

Obtaining KT testing in our cohort did not require review or approval by an infectious disease specialist and was ordered at the discretion of the evaluating rheumatology provider. Cohorts which have described strict institutional regulation of mcfDNA testing, including requirement of infectious disease approval, have reported higher positive testing rates (61%); however, even with tighter regulation, only 16% of positive tests resulted in a new or earlier diagnosis [14]. Our report is the first of its kind to depict testing characteristics of mcfDNA in an entirely outpatient setting; therefore, further studies are needed to determine the reproducibility of our findings and also to assist in identification of which patient features might be associated with a higher pre-test probability for positive testing among similar patient groups. Given our findings, patients with multifocal or poly-organ pathology suspicious for culture-negative endocarditis, mycobacterial infection, or Whipple’s disease mimicking a rheumatic syndrome would be reasonable to consider testing with mNGS if conventional methods are unable to timely secure a diagnosis.

The definition of clinical relevance and impact are not universally defined and vary between cohort studies. In the current series, 6 cases, accounting for 25% of positive mcfDNA results and 4% of overall tests obtained, were defined as positive and clinically relevant. The definition used in our series was intentionally strict in order to determine how often the mcfDNA test identified an infection as the reason for the symptoms which were referred to our division and presumed to be rheumatologic. While this is noticeably lower than reported clinical impacts of mcfDNA in other populations, this is because other studies have used different and broader definitions. For example, Lee and colleagues evaluated 59 mNGS tests and found 28/51 organisms detected were clinically relevant; however, while the definition used included a positive mNGS test confirmed by conventional methods and associated with the primary etiology of illness, it also included, as clinically relevant, confirmed positives that were not the primary reason for hospitalization/acute illness [11]. Similarly, Shishido et al. described a positive clinical impact of mNGS testing on treatment decisions in 43% of cases among 80 patients, 56% of which were immunocompromised [14]. Although positive impact included testing that led to a new diagnosis or earlier diagnosis, it also attributed positive impact to avoidance of invasive procedures, reduction in length of hospital stay, initiation of appropriate antimicrobial therapy, de-escalation/discontinuation of antimicrobial therapy and confirmation of clinical diagnosis [14]. It is notable that the latter two were the primary drivers of clinical benefit in nearly 80% of positive impact results and positive mNGS leading to new diagnosis was only observed in 6 cases [14].

In patients where infection is presumed and empiric treatment initiated, de-escalation of antimicrobial therapy following mNGS testing is a key benefit of mcfDNA technology. Conversely, among patients presenting with known or suspected rheumatic disease, excluding infection prior to initiation or escalation of immunosuppression is paramount. While this study did not pre-assign a definition of positive impact to a negative mNGS test prior to treatment initiation, it is noteworthy and beneficial that 51% with negative mNGS testing initiated or escalated immunosuppression without evidence of subsequent infection observed in the next 3 months, suggesting that infection was not a contributor to the presenting features at the time of testing. While we would not advocate for routine mNGS prior to starting or increasing immunosuppression, in cases where clinical features raise the concern for atypical infection, a negative mNGS result can provide reassurance and is in keeping with other groups reporting calculated sensitivity estimates of 93% and negative predictive values of 81-99% [6, 12].

KT can be negative despite presence of active infection, as noted by five cases where the mNGS testing was negative but an infection was confirmed by alternative methods. The false negative rates in our study are similar to other cohorts [6, 15] but lower than a study evaluating pneumonia in immunocompromised patients where mNGS was negative in 20% of patients where usual care methods identified a causative organism [7]. The low rate of false negative in our cohort may be the result of infrequent utilization of empiric antimicrobial therapy. Four of the five patients in our study required surgical or procedural approaches to confirm the pathogen. Therefore, if infection remains clinically suspected despite mNGS testing, invasive procedures may be required. Three of the patients with negative mNGS were on anti-TNF therapy. While it is not possible to extrapolate based on a limited sample size, further investigations are needed to evaluate any potential correlations.

Although this report is strengthened by the larger sample size in comparison to studies using similar technologies, this study is limited by its single center, retrospective design. Thus, the findings may be less generalizable to other populations. Additionally, data extracted from the medical record regarding reason for testing may be subject to error. Unless explicitly stated by the ordering provider in their documentation, determining if a KT was obtained to assess for FUO, assess an atypical RD presentation, or to assess for RD flare versus infection in the setting of immunosuppression was inferred by the researcher based on the reason for the clinic appointment, patient presentation, and provider assessment and plan documentation.

## Conclusion

For select patients within an outpatient rheumatology practice, mNGS of mcfDNA was useful in distinguishing infection from RD flare or atypical RD presentation. Sequencing of mcfDNA was critical to timely identification and treatment of infection in six patients. For patients without a pathogen identified via mcfDNA sequencing, benefit was provided by reducing suspicion of serious infection prior to initiation or escalation of immunosuppression. To maximize benefit and minimize cost, it will be important to continue investigations to further define the patients who will most benefit from mcfDNA testing in the rheumatology setting.

## Data Availability

All data produced in the present study are available upon reasonable request to the authors

## Acknowledgements

The authors thank Karius® for providing a comprehensive list of all patients for which outpatient testing was completed at our institution during the study dates of interest.

## Author contributions

All authors contributed to at least one of the following manuscript preparation roles: conceptualization AND/OR methodology, data abstraction, formal analysis, data curation AND drafting or reviewing/editing the final draft. As corresponding author, Dr. Jenkins confirms that all authors have provided the final approval of the version to be published, and takes responsibility for the affirmations regarding article submission, the integrity of the data presented, and the statements regarding compliance with institutional review board/Helsinki Declaration requirements.

## Funding

No funding was received for any work related to this study

## Ethics statement

This study was reviewed and approved by the institutional review board, and research was conducted in compliance with Helsinki Declaration requirements.

## Conflict of Interest

None of the authors have conflicts of interest to declare in association with this study.

## Notes

### Competing Interest Statement

The authors have declared no competing interest.

### Funding Statement

This study did not receive any funding

### Author Declarations

IRB of Mayo Clinic waived ethical approval for this work.

